# Retinal vascular trajectories and their associations with systemic health in the Canadian Longitudinal Study on Aging

**DOI:** 10.64898/2026.10.01.26364520

**Authors:** Thomas H Julian, George Moussa, Georgia R Morgan, Tom MacGillivray, Yitian Zhao, Bernard Keavney, Matthew Sperrin, Alejandro F Frangi, Panagiotis I Sergouniotis

## Abstract

Retinal vascular features are proposed biomarkers of systemic health, yet their longitudinal behaviour and relationship to systemic health remain poorly understood. We quantified retinal vascular morphology from serial colour fundus photographs in the Canadian Longitudinal Study on Aging, including 21,562 participants at baseline with good quality images. On average, vascular complexity and density decreased between baseline visit and follow-up 2 (mean interval 5.7 years), whereas tortuosity, arteriolar widths, and venular widths increased. Baseline vascular features were associated with demographic, cardiovascular, metabolic and behavioural traits, whereas altered longitudinal trajectories were associated principally with older age and higher low-density lipoprotein cholesterol. Within-person increases in mean arterial pressure covaried with lower vascular complexity, density and vessel widths. In survival analyses, independently of baseline vascular features, increases in fractal dimension were associated with lower hazard of subsequent all-cause mortality (hazard ratio = 0.92 per standard deviation, 95% confidence interval (CI) = 0.86 - 0.98), whereas arteriolar widening was associated with higher mortality (hazard ratio = 1.09, 95% CI = 1.02 - 1.17). No retinal change metric was associated with major adverse cardiovascular events after false-discovery-rate correction. These findings identify retinal vascular remodelling as a dynamic phenotype associated with systemic health and subsequent mortality, and motivate evaluation of multi-time-point retinal imaging in oculomics.

## Introduction

The retina is the only tissue that permits direct, non-invasive visualization of the microvasculature and central nervous system.^1^ The retinal vasculature in particular has received a great deal of attention in the realm of systemic health modelling due to the association between retinal vascular morphology and future risk of cardiovascular and neurological disorders. The most frequently studied imaging modality in this domain is colour fundus photography, a modality which offers a clear view of retinal arterioles and venules and is widely available in high street optometry.^1–8^

Despite extensive research linking retinal vasculature to systemic health, there is a scarcity of longitudinal studies examining how retinal vessels change with age, altered behaviours, and systemic health factors.^9–13^ Most studies linking retinal vascular anatomy to systemic health have been cross-sectional in nature, limiting our understanding of the extent to which retinal vascular features are stable health biomarkers, or dynamic tissue phenotypes that remodel in response to changing physiology and behaviour.^1,4,5,7,9–11,14–16^ Within the few longitudinal studies that have explored retinal vascular features with respect to health, little attention has been given to time interactions.^9,10^ Moreover, to our knowledge, no study has explored how longitudinal changes in retinal vascular features are associated with subsequent disease risk.

The Canadian Longitudinal Study on Aging (CLSA) dataset offers the opportunity to address this gap because the same participants have been assessed across three rounds of data collection, with each visit featuring colour fundus photography.^17^ This design enables the study of within-person and population-level change in retinal vascular structure over time.^18,19^ It also allows longitudinal retinal vascular morphological changes to be explored as an indicator of subsequent disease risk, enabling assessment of the relationship between the rate of remodeling and future disease events.

In this study, we sought to address three related questions: (1) the extent to which the retinal vasculature changes over time; (2) whether behavioural and systemic health factors are associated with differences in retinal vascular trajectories; and (3) whether interval changes in retinal vascular features are associated with the subsequent hazard of disease, independent of baseline retinal vascular phenotype.

## Results

### Population Characteristics

The eligible study population was 51.7% female, the mean age of participants at baseline was 61.14 (standard deviation [SD] = 9.63, range = 45 to 86), and 94.8% were of self-reported white ethnicity. Representation of non-white ethnic groups was sparse, with the second largest ethnic group (mixed ethnicity) representing just 1.5% of participants, and most others a fraction of a percentage point (precluding meaningful analyses on the role of ethnicity in ophthalmic features).

The total participants with right eye colour fundus photographs satisfying quality control criteria reduced over the duration of the study follow-up - with 21,562 participants at baseline, reducing to 15,666 and 8,312 at follow-up 1 and follow-up 2 respectively. As shown in **Figure 1**, a substantial proportion of the loss to follow-up, particularly at follow-up 2, was due to participants who attended the data collection site visit not undergoing right eye imaging. The reasons for non-capture of right eye images are not reported by the CLSA. Across all participants, the mean baseline to follow-up 1 interval was 2.93 years (SD = 0.29 years, range = 1.30 to 5.96) and the mean follow-up 1 to follow-up 2 interval was 2.79 years (SD = 0.24 years, range = 1.84 to 4.31). Complete demographic characteristics of the cohort are presented in **Supplementary Figure 1**.

**Figure 1:**
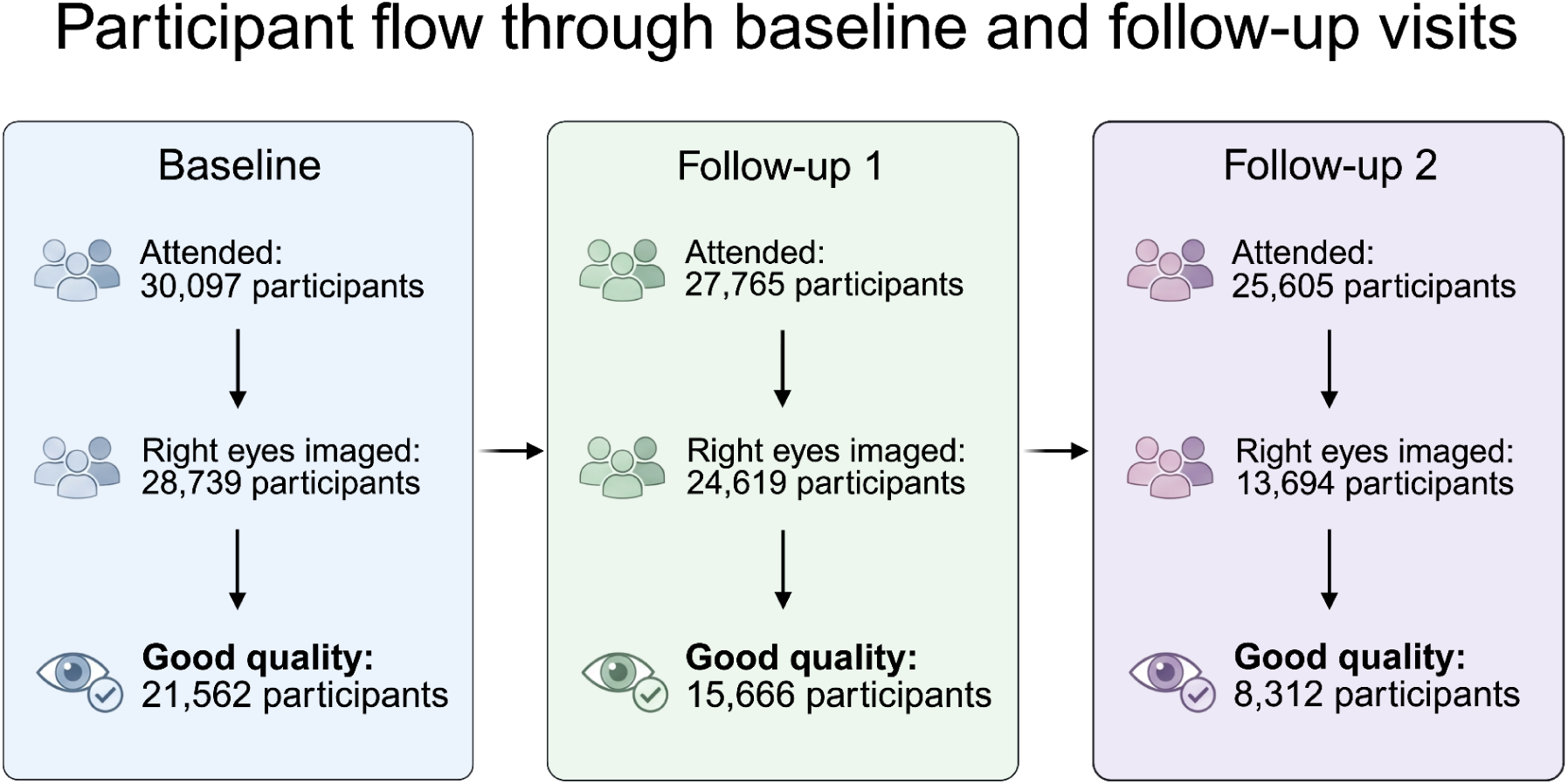
A participant flow diagram across baseline, follow-up 1 and follow-up 2. The three boxes indicate total subjects at baseline, follow-up 1 and follow-up two. In each, the top row is all subjects who attended the study data collection site visit. The second row indicates the number of those subjects who underwent right eye imaging. The bottom row indicates the number of subjects who subsequently passed quality control as described in the Methods.

### Vascular metric change scores are correlated

Retinal vascular metrics included (i) a measure of vessel complexity (fractal dimension); (ii) a measure of the non-smooth appearance of the vessel course (tortuosity) (iii) the ratio between the area of vessels to the area of the whole image (vessel density); and (iv) the average width of venules and arterioles. Baseline to follow-up 2 change scores in retinal vascular metric change values were calculated (distributions of percentage change in values are shown in **Figure 2**). To determine whether the selected retinal parameters capture overlapping or distinct aspects of longitudinal microvascular aging, we evaluated the correlations among their respective change scores. All vascular metric change scores (represented as Δ, and measured from baseline to follow-up 2) were correlated at a level exceeding multiple testing correction (**Figure 3**). The most closely related metrics were Δ fractal dimension and Δ vascular density (*r* = 0.87), followed by Δ average arteriolar and Δ venular width (*r* = 0.72). Moderate correlations were observed for all other trait combinations. These results show that retinal vascular metrics are correlated, but are sufficiently distinct to warrant their individual inclusion in the analysis. The complete results are presented in **Supplementary Table S1**. All correlations were statistically significant (*P* < 0.001), and remained significant after correction for multiple testing using FDR.

**Figure 2:**
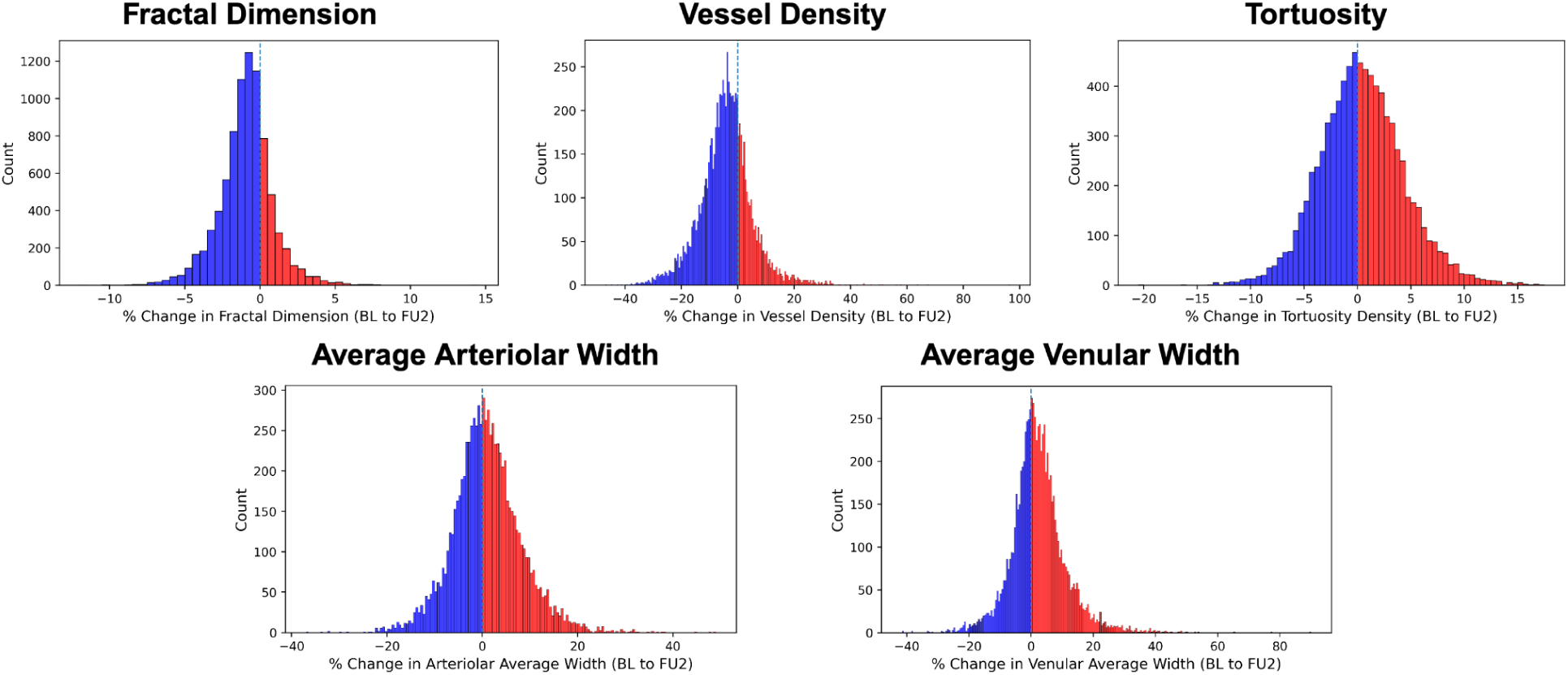
The distribution of subject percentage change in vessel metric values from baseline to follow-up 2. Red coloured columns represent increased values, whilst blue colouring indicates decreased values. As shown, whilst there were prevailing directions of change, every metric showed change in both directions across the cohort.

**Figure 3:**
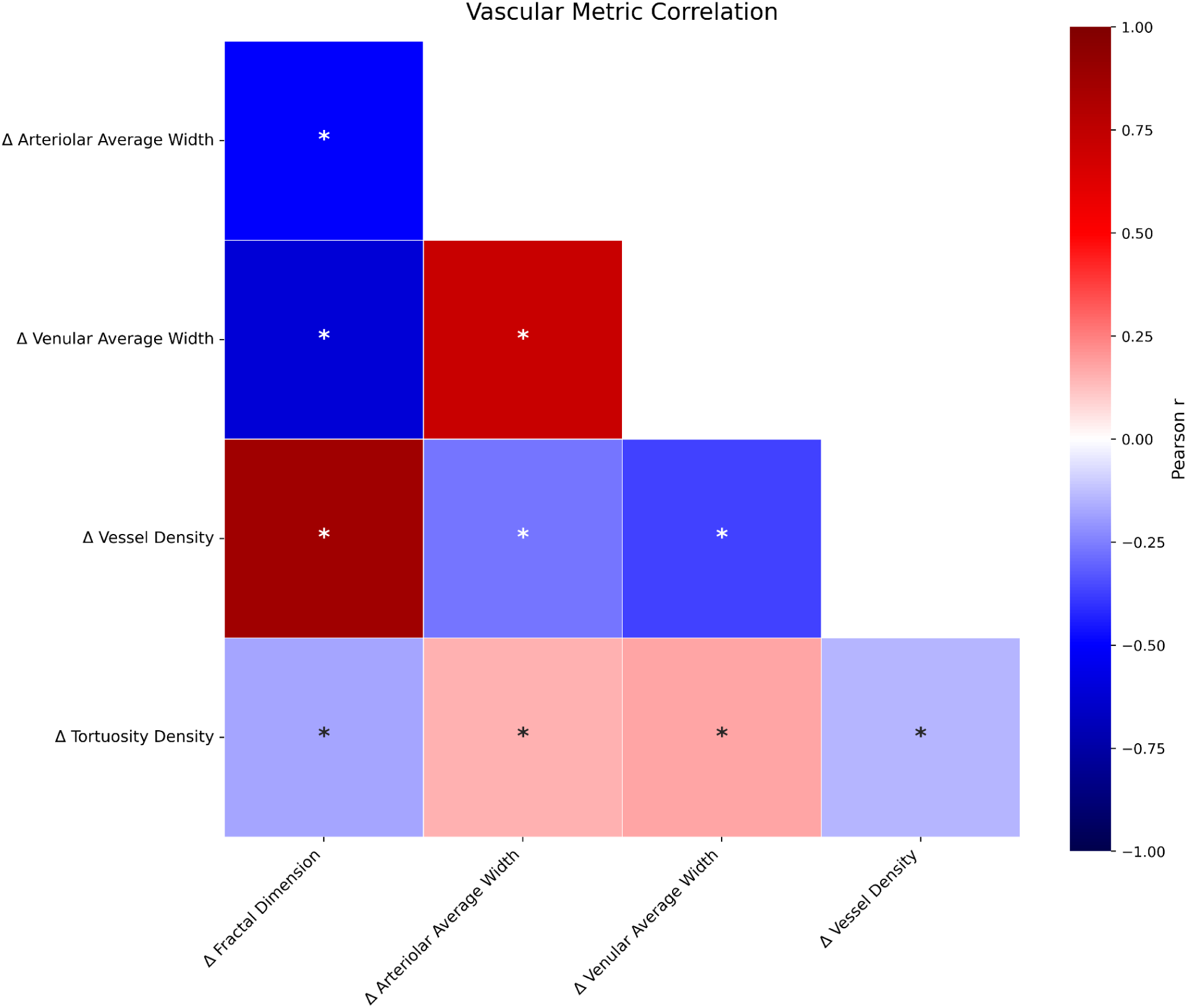
A correlation plot, which illustrates the baseline-follow-up 2 change score correlations (indicated by Δ). An asterisk (*) indicates that the result was significant after FDR correction for multiplicity. P values are two-tailed.

### The evolution of the retinal vasculature with age

From baseline to follow-up 2 (mean interval = 5.7 years, SD = 0.35 years), retinal vascular measures showed modest changes, with directionally consistent results between baseline to follow-up 1 and baseline to follow-up 2. Although the direction of vascular change was variable between participants (i.e. in some individuals an individual metric increased, and in some it decreased), each metric showed a statistically significant mean change over time in linear mixed models. Among 8,312 participants with paired baseline to follow-up 2 measurements, over the duration of the study, measures of vascular complexity decreased, with fractal dimension decreasing by a mean of 0.93% (SD = 1.82%; range = −11.52% to 14.15%), and vessel density decreasing by a mean of 4.17% (SD = 9.55%; range = −46.85% to 96.10%). Meanwhile, overall, tortuosity density increased slightly, by a mean of 0.42% (SD = 3.97%; range = −20.42% to 17.14%) as did vessel calibres, with arteriolar average width increasing by a mean of 0.98% (SD = 7.29%; range = −36.57% to 48.68%) and venular average width increasing by a mean of 2.30% (SD = 8.88%; range = −41.36% to 89.88%). The change score distributions are illustrated graphically in **Figure 2**.

In adjusted random-intercept mixed-effects models, the conditional fixed effect of elapsed time indicated annual decreases of 0.07 SD in fractal dimension and 0.06 SD in vessel density, together with annual increases of 0.009 SD in tortuosity, 0.01 SD in arteriolar average width and 0.03 SD in venular average width.

The conditional model estimates (linear mixed models) and unadjusted mean percentage changes (as shown in **Figure 2**) calculated in the paired follow-up samples differ in their assumptions and method, but both analyses showed concordant directions of change: decreasing retinal vascular complexity and density, increasing tortuosity, and widening of arteries and, more prominently, venules (**Table 1**). Interestingly, while arteriolar width was shown to increase with time, at the baseline visit retinal arterioles were shown to be significantly narrower with older age (**Table 2**).

**Table 1:** The percent change in vascular metric values over the course of follow up, and the conditional rate of change after adjusting for age, sex, health behaviours and cardiovascular health metrics. The conditional rate of change is expressed as the number of standard deviations (SD) of change per year. The conditional rate change estimates describe the expected annual changes for a female participant with the continuous characteristics included in time-interaction terms as described in the methods. Retinal outcomes were standardised separately for each metric and eye using the mean and SD of all complete-case observations across the three visits.

| Metric | Mean BL–FU1 change (%) | Mean BL–FU2 change (%) | Conditional rate of change per year (SD-change) |
| --- | --- | --- | --- |
| Fractal dimension | -0.54 | -0.93 | -0.07 |
| Vessel density | -2.32 | -4.17 | -0.06 |
| Tortuosity | +0.25 | +0.42 | +0.009 |
| Arteriolar average width | +0.86 | +0.98 | +0.01 |
| Venular average width | +1.46 | +2.30 | +0.03 |

**Table 2:** A summary of the results of mixed linear modelling. Additional baseline associations are reported in the supplementary content. Bold, blue text highlights that the FDR correct *P* value (FDR *P*) is < 0.05. N = 33,557 observations from 17,288 participants, except arteriolar average width (33,556 observations). MAP, mean arterial pressure; BMI, body mass index; PASE, Physical Activity Scale for the Elderly; LDL, low-density lipoprotein; HDL, high-density lipoprotein; HbA1c, glycated haemoglobin. A small number of *P* values are smaller than numerical precision of the python programme and are therefore displayed as *P* < 0.001.

| A. Differences in annual retinal change (exposure x elapsed-time interactions) |  |  |  |  |  |
| --- | --- | --- | --- | --- | --- |
| Predictor (unit) | Fractal dimension | Vessel density | Tortuosity density | Arteriolar average width | Venular average width |
| Sex (male vs female) | -0.002 (-0.009, 0.005)<br>FDR $P = 0.65$ | 0.001 (-0.005, 0.007)<br>FDR $P = 0.65$ | 0.008 (0.0004, 0.02)<br>FDR $P = 0.07$ | 0.008 (0.0008, 0.02)<br>FDR $P = 0.07$ | 0.009 (0.0005, 0.02)<br>FDR $P = 0.07$ |
| Participant-mean MAP (per 1 SD) | -0.001 (-0.004, 0.002)<br>FDR $P = 0.66$ | -0.001 (-0.004, 0.002)<br>FDR $P = 0.66$ | -0.002 (-0.006, 0.002)<br>FDR $P = 0.66$ | 0.0002 (-0.003, 0.004)<br>FDR $P = 0.92$ | -0.002 (-0.006, 0.002)<br>FDR $P = 0.66$ |
| Participant-mean BMI (per 1 SD) | 0.002 (−0.002, 0.006)<br>FDR $P = 0.33$ | 0.003 (−0.0002, 0.006)<br>FDR $P = 0.29$ | 0.003 (−0.001, 0.007)<br>FDR $P = 0.29$ | 0.003 (−0.0009, 0.006)<br>FDR $P = 0.29$ | 0.0005 (−0.004, 0.005)<br>FDR $P = 0.82$ |
| Baseline age (per 5 years) | **−0.008 (−0.01, −0.006)**<br>FDR $P = 1E-16$ | **−0.007 (−0.008, −0.005)**<br>FDR $P = 5E-16$ | 0.0002 (−0.002, 0.002)<br>FDR $P = 0.84$ | **0.004 (0.002, 0.005)**<br>FDR $P = 0.0002$ | **0.005 (0.003, 0.007)**<br>FDR $P = 0.00003$ |
| Participant-mean alcohol intake (per 50 g/week) | −0.0008 (−0.003, 0.001)<br>FDR $P = 0.54$ | −0.0006 (−0.002, 0.001)<br>FDR $P = 0.54$ | 0.001 (−0.001, 0.003)<br>FDR $P = 0.54$ | −0.0002 (−0.002, 0.002)<br>FDR $P = 0.82$ | 0.001 (−0.0009, 0.004)<br>FDR $P = 0.54$ |
| Baseline smoking exposure (per 10 pack-years) | −0.0009 (−0.003, 0.001)<br>FDR $P = 0.68$ | −0.001 (−0.003, 0.0004)<br>FDR $P = 0.38$ | 0.0006 (−0.002, 0.003)<br>FDR $P = 0.82$ | 0.002 (−0.0006, 0.004)<br>FDR $P = 0.38$ | −0.0002 (−0.003, 0.002)<br>FDR $P = 0.89$ |
| Baseline physical activity (per 1 SD PASE) | 0.002 (−0.001, 0.005)<br>FDR $P = 0.61$ | 0.0006 (−0.002, 0.003)<br>FDR $P = 0.71$ | −0.0007 (−0.004, 0.003)<br>FDR $P = 0.71$ | −0.003 (−0.007, −0.0002)<br>FDR $P = 0.20$ | −0.002 (−0.006, 0.002)<br>FDR $P = 0.62$ |
| Baseline LDL cholesterol (per 1 SD) | **−0.004 (−0.007, −0.0008)**<br>FDR $P = 0.03$ | **−0.005 (−0.008, −0.002)**<br>FDR $P = 0.001$ | −0.0009 (−0.004, 0.003)<br>FDR $P = 0.74$ | 0.002 (−0.001, 0.005)<br>FDR $P = 0.35$ | 0.0006 (−0.003, 0.004)<br>FDR $P = 0.74$ |
| Baseline HDL cholesterol (per 1 SD) | −0.0009 (−0.004, 0.003)<br>FDR $P = 0.79$ | 0.001 (−0.002, 0.005)<br>FDR $P = 0.76$ | 0.002 (−0.002, 0.006)<br>FDR $P = 0.76$ | 0.004 (−0.00007, 0.007)<br>FDR $P = 0.27$ | 0.0006 (−0.004, 0.005)<br>FDR $P = 0.80$ |
| Baseline HbA1c (per 1 SD) | −0.002 (−0.005, 0.002)<br>FDR $P = 0.64$ | −0.002 (−0.005, 0.0008)<br>FDR $P = 0.64$ | −0.00003 (−0.004, 0.004)<br>FDR $P = 0.99$ | 0.001 (−0.002, 0.005)<br>FDR $P = 0.64$ | 0.002 (−0.002, 0.006)<br>FDR $P = 0.64$ |
| **B. Concurrent within-person associations** |  |  |  |  |  |
| **Predictor (unit)** | **Fractal dimension** | **Vessel density** | **Tortuosity density** | **Arteriolar average width** | **Venular average width** |
| Within-person MAP deviation (per 1 SD) | **−0.03 (−0.03, −0.02)**<br>FDR $P = 6E-21$ | **−0.05 (−0.05, −0.04)**<br>FDR $P = 5E-71$ | −0.001 (−0.008, 0.005)<br>FDR $P = 0.68$ | **−0.04 (−0.05, −0.04)**<br>FDR $P = 1E-47$ | **−0.02 (−0.03, −0.01)**<br>FDR $P = 1E-7$ |
| Within-person BMI deviation (per 1 SD) | 0.003 (−0.003, 0.009)<br>FDR $P = 0.47$ | 0.003 (−0.002, 0.008)<br>FDR $P = 0.47$ | 0.002 (−0.004, 0.009)<br>FDR $P = 0.54$ | **−0.01 (−0.02, −0.007)**<br>FDR $P = 0.00009$ | 0.003 (−0.004, 0.01)<br>FDR $P = 0.47$ |
| Within-person alcohol deviation (per 50 g/week) | −0.00008 (−0.007, 0.007)<br>FDR $P = 0.98$ | −0.002 (−0.008, 0.004)<br>FDR $P = 0.76$ | −0.004 (−0.01, 0.004)<br>FDR $P = 0.76$ | −0.002 (−0.009, 0.005)<br>FDR $P = 0.76$ | 0.004 (−0.004, 0.01)<br>FDR $P = 0.76$ |
| **C. Annual retinal change for the reference profile** |  |  |  |  |  |
| **Predictor (unit)** | **Fractal dimension** | **Vessel density** | **Tortuosity density** | **Arteriolar average width** | **Venular average width** |
| Elapsed time (per year) | **−0.07 (−0.08, −0.07)**<br>FDR $P = 2E-233$ | **−0.06 (−0.07, −0.06)**<br>FDR $P = 3E-230$ | **0.009 (0.004, 0.01)**<br>FDR $P = 0.0006$ | **0.01 (0.006, 0.01)**<br>FDR $P = 0.00001$ | **0.03 (0.02, 0.03)**<br>FDR $P = 3E-23$ |

### Longitudinal associations between retinal vascular traits, health behaviours, and systemic traits

Linear mixed-effects models were used to investigate associations between retinal vascular features and systemic health characteristics using available measurements across the three CLSA study visits. Retinal vascular metrics were standardised (z-scored), and thus the coefficients represent the number of standard deviations per unit change in the independent variable. Models featured baseline age, sex, smoking pack-years, Physical Activity Scale for the Elderly (PASE) score^20^, HDL cholesterol, LDL cholesterol, HbA1c, and alcohol consumption, and the interaction of these variables with time. Derived variables, any pre-processing applied, and their definitions are described in the **Methods**. The models were designed to distinguish between baseline trait associations (ie the association between traits and baseline vascular values), concurrent within-person variation (ie how changes in a particular trait covary with retinal vascular changes), and the effect of traits on retinal vascular trajectories (assessed with time interactions).^18,19^

After FDR correction, there was evidence that baseline age and serum LDL cholesterol modified the trajectories of retinal vascular features. Each additional five years of baseline age was associated with a greater annual decrease in fractal dimension (β = −0.008 SD/year, 95% CI−0.01 to −0.006, FDR-adjusted *P* = 1.20 × 10^-16^) and vessel density (β = −0.007 SD/year, 95% CI −0.008 to −0.005, FDR-adjusted *P* = 5.35 × 10^-16^), and a greater annual increase in average arteriolar width (β = 0.004 SD/year, 95% CI 0.002 to 0.005, FDR-adjusted *P* = 1.94 × 10^-4^) and average venular width (β = 0.005 SD/year, 95% CI 0.003 to 0.007, FDR-adjusted *P* = 3.13 × 10^-5^). Per 1-SD higher baseline LDL cholesterol, fractal dimension and vessel density declined faster by 0.004 SD/year (95% CI −0.007 to −0.0008, FDR-adjusted *P* = 0.04) and 0.005 SD/year (95% CI −0.008 to −0.002, FDR-adjusted *P* = 0.001), respectively. Physical activity levels (PASE) were nominally significantly related to rate of change in average arteriolar width (*P* = 0.04), but this did not persist after multiple testing correction (FDR-adjusted *P* = 0.20). Although sex was nominally related to rate of change in average venular width, arteriolar width, and tortuosity, this did not pass multiple testing corrections. None of the other tested characteristics modified the trajectory of any retinal feature after FDR correction.

Significant within-person (deviation score) associations were identified for mean arterial pressure (MAP). At visits when a participant’s MAP was 1 SD above their own mean, fractal dimension (β = −0.03 SD, 95% CI −0.03 to −0.02, FDR-adjusted *P* = 5.69 × 10^-21^), vessel density (β = −0.05 SD, 95% CI −0.05 to −0.04, FDR-adjusted *P* = 4.80 × 10^-71^), average arteriolar width (β = −0.04 SD, 95% CI −0.05 to −0.04, FDR-adjusted *P* = 1.13 × 10 ^-47^) and average venular width (β = −0.02 SD, 95% CI −0.02 to −0.01, FDR-adjusted *P* = 1.17 × 10^-7^) were lower. Meanwhile, tortuosity was not associated with within-person MAP variation (FDR-adjusted *P* = 0.68). A 1-SD within-person increase in body mass index (BMI) was associated only with lower average arteriolar width (β = −0.01 SD, 95% CI −0.02 to −0.007, FDR-adjusted *P* = 9.41 × 10^-5^), but with no other vascular metrics. Within-person variation in alcohol consumption was not associated with any metric.

At baseline, broader associations were present for numerous traits. Age, sex and participant-mean MAP were each associated with all five retinal features. Whilst there were retinal vascular associations with baseline BMI, alcohol consumption, smoking, physical activity, LDL cholesterol, HDL cholesterol and HbA1c, these were not universally significant across retinal vascular metrics after multiple testing correction. Complete results, including null results, are reported in **Table 2**, and are expanded in **Supplementary Table S2**.

### Baseline to follow-up one vascular metric change scores and future health

With a view to exploring the extent to which absolute retinal vascular change between baseline and follow up 1 was a predictor of future ill health, we utilised Cox proportional hazards models adjusted for sex and baseline vascular metric. The model used attained age as the time scale, providing control for age effects.^21^ The events modelled were three-point major adverse cardiovascular events [MACE] and all-cause death.^22^ The results of Cox proportional hazards modelling are presented in **Table 3**, and with additional information (coefficients and follow-up times) presented in **Supplementary Tables S4 & S5**.

**Table 3:** The results of Cox proportional hazards. This table shows the hazard ratio, confidence intervals (CI) and p values for the change values for each retinal vascular metric adjusted for sex and baseline metric values. MACE = major adverse cardiovascular events, SD = standard deviation, N = sample size, events = total events during follow up, FDR = false discovery correction. In this cohort, there were 177 MACE events and 837 all-cause mortalities.

| Event | Retinal Metric | HR | Lower 95% CI | Upper 95% CI | N | Events | $P$ Value | FDR $P$ Value |
| --- | --- | --- | --- | --- | --- | --- | --- | --- |
| Death | Change in Fractal Dimension (per SD) | 0.92 | 0.86 | 0.98 | 15575 | 837 | 0.01 | 0.03 |
| Death | Change in Vessel Density (per SD) | 0.94 | 0.88 | 1.00 | 15575 | 837 | 0.05 | 0.09 |
| Death | Change in Tortuosity Density (per SD) | 1.06 | 0.99 | 1.13 | 15575 | 837 | 0.08 | 0.10 |
| Death | Change in arteriolar Average Width (per SD) | 1.09 | 1.02 | 1.17 | 15574 | 837 | 0.01 | 0.04 |
| Death | Change in venular Average Width (per SD) | 1.03 | 0.97 | 1.10 | 15575 | 837 | 0.36 | 0.36 |
| MACE | Change in Fractal Dimension (per SD) | 0.87 | 0.76 | 1 | 14424 | 177 | 0.05 | 0.27 |
| MACE | Change in Vessel Density (per SD) | 0.91 | 0.79 | 1.05 | 14424 | 177 | 0.21 | 0.36 |
| MACE | Change in Tortuosity Density (per SD) | 1.06 | 0.92 | 1.23 | 14424 | 177 | 0.4 | 0.4 |
| MACE | Change in arteriolar Average Width (per SD) | 1.12 | 0.97 | 1.31 | 14423 | 177 | 0.13 | 0.32 |
| MACE | Change in venular Average Width (per SD) | 1.07 | 0.92 | 1.24 | 14424 | 177 | 0.37 | 0.4 |

After FDR correction, a one SD increase in fractal dimension between baseline and follow-up one was associated with 8% less hazard of death between follow-up 1 and 2 (FDR-adjusted *P* = 0.03). Meanwhile, a one standard deviation increase in average arteriolar width was associated with 9% higher hazard of death. Change in vessel density was of borderline nominal significance in relation to decreased risk of death (*P* = 0.05), but this did not exceed multiple testing correction (FDR-adjusted *P* = 0.09). Similarly, although increasing fractal dimension was of borderline nominal significance in relation to decreasing hazard of MACE (*P* = 0.05), this did not exceed multiple testing correction (FDR-adjusted *P* = 0.27). No other retinal vessel metrics were associated with hazard of death or MACE.

## Discussion

This longitudinal analysis demonstrated that retinal vascular metrics evolve with time, showing moderate variability over the course of follow-up in the CLSA study after adjustment for a number of cardiovascular health-related traits. Whilst cross-sectional relationships between retinal vascular traits and age have been extensively reported, changes to the retinal vasculature with time have received little attention in longitudinal studies outside of ocular disease specific research.^7,23^ In this study we showed that over time, the complexity and density of the retinal vascular system decreases, tortuosity increases, and the retinal vasculature becomes more dilated. Interestingly, at baseline, our study also indicated that retinal arterioles were narrower with older age - a finding seemingly at initial contradiction with our longitudinal results. The relative magnitude of retinal vascular changes was in most cases modest, with the highest level of variation evident for vessel density, which decreased on average by 4.17% over a mean follow-up duration of 5.7 years (**Table 1)**. Our findings are largely in keeping with previous studies reporting the relationship between age and retinal vascular metrics cross-sectionally, which have shown decreased fractal dimension and vessel density in older participants, increased venular diameter, and increased tortuosity.^10,15,24–26^ However, our longitudinal observations with respect to arteriolar width are in contrast with the findings of the cross-sectional literature, which has largely reported decreased arteriolar width with older age.^11,26–28^ We postulate that the apparent contradiction, whereby baseline cross-sectional narrowing contrasts with longitudinal widening, reflects distinct stages of vascular aging. Specifically, it may be that cross-sectional narrowing at baseline captures cumulative early-mid life arteriosclerosis and structural lumen loss, whereas longitudinal widening over the follow-up period reflects endothelial dysfunction-related dilatation in later life.^11,29^ The interpretation that our findings could reflect staged vascular aging is supported by our finding that each additional five years of baseline age predicted steeper annual decreases in vascular complexity and density, alongside greater increases in average arteriolar and venular widths.

Consistent with the wider literature, our study indicated that the baseline retinal vascular features were associated with health behaviours, including baseline physical activity levels, baseline pack years of smoking, and longitudinal alcohol consumption.^6,9,10,12^ Conversely, neither baseline pack years of smoking nor alcohol consumption (including within-person changes to alcohol consumption over the course of the study) were associated with the trajectory of retinal vascular features over time in a model adjusted for other key covariates. Whilst baseline physical activity levels were nominally significantly associated with altered trajectory of average arteriolar width, this did not persist after correction for multiplicity, and there was no evidence that other vascular trajectories were related to PASE score.

All cardiovascular-health related traits tested, including BMI, MAP, and blood markers (LDL, HDL and HbA1c) were associated with retinal baseline vascular features. These findings are in line with previous literature which has indicated that the retina is reflective of adverse cardiovascular health.^1,4,5,13,14,30–32^

Our study showed that within-person increases in MAP were also associated with lower fractal dimension, lower vessel density, lower average arteriolar width and lower average venular width; but that between-person MAP was not associated with altered retinal vascular trajectories. Therefore, MAP is associated with retinal vascular phenotypes, including within-person covariation, but average MAP does not appear to determine the underlying rate of retinal vascular change. There are two possible interpretations of this observation: (a) MAP dynamically impacts the retinal vasculature, but these changes are reversible (therefore affecting covariation, but not long term retinal vascular trajectory) and/or (b) elevated MAP does alter retinal vascular trajectory, but pathological changes in this cohort occurred before entry to the study (given the age distribution of CLSA participants). The existing literature, which features numerous studies on the effects of antihypertensives on the retinal vasculature, supports the assertion that hypertension-associated retinal vascular phenotypes are responsive to improved blood pressure, but the full extent of reversibility to baseline features pre-hypertension is unknown.^33–35^ As our study did not model the impact of medication use on the retinal vasculature, we are unable to disentangle pharmacological retinal vascular changes from MAP-only retinal vascular variation.

Higher baseline blood LDL cholesterol was also associated with the trajectory of retinal vascular complexity, specifically a greater annual decrease in fractal dimension and vessel density.

Baseline HDL cholesterol, meanwhile, was not associated with altered retinal vascular trajectory. To our knowledge, whilst there is a wealth of cross-sectional evidence suggesting a relationship between circulating lipoproteins and retinal vascular traits, there is a paucity of longitudinal research and evidence of altered trajectories. While there are reports of vessel calibre being longitudinally associated with serum lipoproteins, evidence around markers of retinal vascular complexity is lacking.^10,36^ With that said, the present study’s findings indicating that LDL cholesterol is associated with reduced complexity of the retinal vascular system is highly biologically plausible given the impact of lipid physiology on systemic vascular health ^37^, and evidence that poor cardiovascular health is associated with reduced retinal vascular complexity.^14^

With respect to BMI, our within-person analysis suggested that increased body mass was associated with lower average arteriolar width, but no other retinal vascular measures.

Counter-intuitively, with adjustment for other cardiovascular disease related covariates, BMI was not associated with altered retinal vascular trajectories. Whilst there has been previous longitudinal research indicating that BMI may alter the retinal vasculature in children and adults, the relevant studies do not appear to have decomposed BMI into between- and within-person variation, or explored how such relationships interact with time.^10,32^

Finally, our study explored how baseline to follow-up one retinal vascular change scores related to future hazard of all-cause death and three-point MACE. We found that an interval-increase in fractal dimension was associated with reduced hazard of all-cause death (8% per standard deviation), and increased arteriolar average width was associated with increased hazard of death (9% per standard deviation). Whilst fractal dimension was of borderline nominal significance in relation to MACE hazard, this result did not exceed multiple testing correction. No other vascular change metrics were associated with MACE or all-cause death. Importantly, whilst there were a large number of death events (N=837), there were just 177 MACE events during follow-up. We are therefore unable to exclude the possibility that our null MACE results were a product of insufficient power. Our results with respect to all-cause death suggest that interval changes in retinal vascular metrics are of significance with respect to future systemic health outcomes. This is a notable finding for the field of oculomics, given that models developed this far utilise single-instance ophthalmic imaging events to model future disease risk.^1,16,38,39^

This study has several limitations. Our study relies on self-reported disease events during interviews with volunteers, their friends, and their relatives which introduces risk of reporting errors. Further, the study featured three visits inclusive of colour fundus photography, with substantial attrition over time. The methodology utilised is posed to measure association between systemic health behaviours and traits, but is not positioned to interrogate causality. Finally, intra-subject variation in vascular measures are influenced by both real anatomical changes and technical factors relating to image quality and image centering. Whilst we mitigated this to the extent possible by utilising only good quality images, it is likely that vascular feature variability estimates are influenced by a combination of both technical factors and true morphological change.

## Conclusion

In this longitudinal population study, retinal vascular features changed systematically over time, with their trajectories varying according to baseline age and LDL cholesterol. Specific interval changes, namely increasing fractal dimension and arteriolar widening, were associated with subsequent all-cause mortality independently of their baseline value. These findings extend oculomics beyond conventional single-time-point analyses by indicating that temporal variation in retinal vascular morphology may provide information about systemic health not captured by baseline morphology alone. Therefore, our analysis suggests that head-to-head comparison of models using single time point and temporally encoded image sequences could be a valuable route for future research. Given that retinal imaging is already routinely repeated over several years in community optometry and secondary eye care, longitudinal phenotypes could be exploited without altering the current workflows implemented in routine eyecare. In summary, this study suggests that the retinal vasculature should be considered not only as a static marker, but also as a dynamic phenotype whose remodelling may provide a complementary perspective on systemic health and future risk.

## Methods

### Data source

Data were obtained from the Canadian Longitudinal Study on Aging (CLSA), a longitudinal study which recruited English or French speaking individuals aged 45-85 years.^17^ The CLSA study was reviewed by twelve Canadian research ethics boards as previously described.^40^ The present study was exempted from requiring local ethical approval by the University of Manchester Research Ethics Team (Reference: 2024-21253-36907) and used the “comprehensive cohort” of the CLSA (the subset of CLSA participants who underwent a range of physical assessments and in person interviews). The comprehensive cohort consisted of 30,097 participants randomly selected from within 25–50 km of eleven data collection sites (DCSs) in seven provinces. These participants were interviewed in person, took part in in-depth physical assessments at DCSs, provided blood samples, and underwent colour fundus photography. Macula-centred colour fundus photographs were captured using a Topcon TRC-nw8 Retinal Camera (45°) in a darkened room without pharmacological mydriasis. The device remained consistent over the course of follow-up. The right eye was imaged, followed by the left eye. For the purpose of this study, right eye images were retained for analysis - with the intention that we maximise sample size (which would be reduced if requiring both eyes to be present in a bilateral analysis), and limit multiplicity (which would be escalated by undertaking the analysis in each eye). This approach has been widely used in previous studies of ophthalmic phenotypes. ^5,9,41,42^ Deaths, and cause of death, were ascertained via a decedent questionnaire that was administered to a close relative or friend after a participant died. Disease events were ascertained via the structured interviews at each follow-up. Our study was conducted using data collected at baseline, follow-up 1 and follow-up 2.

### Retinal Vascular Metrics

Retinal vascular features were derived from colour fundus photographs using AutoMorph, a widely used retinal vascular segmentation and quality control model that has been demonstrated to possess multi-vendor validity.^43^ Retinal vascular metrics utilised in this study included (i) a measure of vessel complexity (fractal dimension / Minkowski–Bouligand dimension); (ii) a measure of the non-smooth appearance of the vessel course (tortuosity, calculated as tortuosity density^44^) (iii) the ratio between the area of vessels to the area of the whole image (vessel density); and (iv) the average width of venules and arterioles. For vascular widths, which are calculated in µm by AutoMorph, pixel resolution was assumed to be the Automorph default for Topcon devices (0.008). As there is variability between Topcon device models and their pixel resolution (e.g. for a DRI Triton vs the camera used in this study), we do not assume the width metrics to be exactly expressed in µm. Notably, the parameter of interest in this study is vessel trajectory rather than absolute values, and therefore the unit scale is not critical.

### Quality Control

Colour fundus photograph quality control was conducted using AutoMorph. Images established to be of “Good” quality were retained for analysis.^43^ Good quality was defined on the basis of image illumination, artifacts, and the diagnosability of the general eye diseases in the AutoMorph methodology. Following calculation of retinal vascular metrics, it was noted that there was a subset of outliers for average venular and arteriolar width evident in the metric histograms (**Supplementary Figures S1-S2**). Accordingly, images with an average arteriolar or venular width value beyond the visualised distribution of the non-outlier population (40 units or less) were excluded from the analysis. Following exclusion, a random sample of 100 excluded images were reviewed by an Ophthalmologist (T.H.J), with 100% of retinal images identified to have major segmentation discontinuities and/or hallucinated vessels (typically short and thin stretches of hallucinated vessels). Conversely, a random sample of 100 included images featured no such hallucinations - suggesting a low frequency of such hallucinations in the wider dataset. Participants were required to have at least a good quality baseline right eye image in order to be considered for analysis.

### Clinical and behavioural variables

Body mass index (BMI) was calculated as body weight in kilograms divided by height in metres squared. Mean arterial pressure (MAP, defined as the time-averaged pressure in peripheral arteries over a cardiac cycle) was calculated as diastolic blood pressure plus one-third of the difference between systolic and diastolic blood pressure.^45^ BMI and MAP were calculated at each available study visit.

In order to assess dose-response to smoking, smoking exposure was expressed as estimated pack-years using a previously described method.^46^

Physical activity was quantified using a total Physical Activity Scale for the Elderly (PASE) score. The PASE score has been demonstrated to offer a representative picture of physical activity in older adults, and has been validated in numerous studies.^47^ The physical activity domains captured in the CLSA interviews were wide-ranging including walking; light, moderate, and strenuous sport and activity; housework; caring for another person; and work/volunteering. The amount of time spent in activities is collected (i.e. duration and frequency) for all except household activities which are dichotomously recorded (yes/no). These responses, along with the predetermined weights for each activity, were used to calculate a total PASE score. The method for calculating PASE score has been described in previous publications.^20,47^ A higher PASE score indicates greater physical activity levels. The first instance of physical activity data were collected during the “maintaining contact questionnaire” via telephone interviews during the early phase of the longitudinal study, and it therefore treated as a baseline variable.

The CLSA collected alcohol consumption data in the form of how many “standard” drinks an individual consumed during typical weekdays and weekends over the course of the last 12 months. A Canadian standard drink is equivalent to 13.45 grams of pure alcohol.^48^ Accordingly, the total standard drinks across all alcohol domains during weekdays and weekends were summed to calculate total grams of ethanol consumed per week. Alcohol consumption data were available across all three CLSA visits.

Additional covariates included age (in years), sex, baseline low density lipoprotein (LDL) cholesterol (mmol/L), baseline high density lipoprotein (HDL) cholesterol (mmol/L) and baseline HbA1c (%). The CLSA does not provide HbA1c values below 4.2%, with all values assigned a coding of “-2222”. In order to facilitate analysis, all lower-limit coded results were assigned a value of 4.2%.

For all data, missing, refused and “don’t know/no answer” codes were converted to missing values according to the coding scheme of each variable. For disease events and the associated date of occurrence, those who refused to answer or answered “don’t know” were excluded from the analysis.

### Correlation between vascular traits

The correlation between retinal vascular change metrics was calculated using Pearson correlation (SciPy, Python) given the normally distributed nature of the variable.^49^

### Longitudinal retinal vascular modelling

The modelling of longitudinal data warrants the use of statistical methods capable of regression analysis of correlated data, given that an individual’s traits across visits are not independent.

Linear mixed effects models are an extension of simple regression models, where the clustering of the repeated measurements within individuals is accounted for through estimation of random effects, and allowing correlated errors.^19^ Accordingly, linear mixed effects models were used in this study to explore the relationship between independent variables of demographic, systemic health, and health behaviour characteristics in relation to retinal vascular features over all three data collection site visits.

Models were fitted separately for each retinal vascular metric. Retinal vascular outcomes were standardised to z-scores. As such, model coefficients represent differences in the retinal vascular outcome in standard deviation units. For all analyses, participants’ visits with data missingness were excluded. Similarly, independent variables were standardised, mean-centred, and expressed in clinically meaningful units as appropriate. This process was undertaken to improve the interpretability of model coefficients.

For repeatedly measured exposures, MAP, BMI, and alcohol consumption were decomposed into between-person (participant mean) and within-person (otherwise known as “deviation score”) components using a hybrid model.^18,50^ The between-person component was defined as each participant’s mean exposure across their available study visits, while the within-person component was defined as the deviation of the exposure at each visit from that participant’s mean exposure. The deviation score / within person metric reflects the differences between the individual observations and the individual mean value and facilitates analysis of vascular metrics and subject health and behaviour variation over the course of the study co-vary. The between subject analysis serves to demonstrate the relationship between the mean value of the particular independent variable for each subject and the vascular metric. This approach enables associations due to differences in average exposure between participants to be distinguished from associations between within-person changes in exposure and concurrent retinal vascular variation. As smoking pack years are cumulative with time and cannot decrease in subsequent visits, deviation scores were not calculated. PASE scores are recent activity estimates taken to proxy for typical activity, but likely to be vulnerable to substantial within person variation, and therefore were also not included in deviation score analyses.

Between-person MAP and BMI were standardised using their distributions across participants. Within-person MAP and BMI components were standardised using the distribution of within-person deviations among participants with at least two observations; participants with only one available observation had a within-person deviation of zero and did not contribute to estimation of the within-person standard deviation. Smoking and alcohol consumption were zero-inflated traits and were therefore not z-standardised. Alcohol consumption was expressed per 50g of ethanol per week, with the between-person component centred on the study mean and the within-person component expressed as the deviation from each participant’s mean consumption. Smoking exposure was mean-centred and expressed per ten pack-years. Baseline age was mean-centred and expressed per five-year increment. PASE score, baseline LDL cholesterol, HDL cholesterol, and HbA1c were z-standardised.

Each model included time since baseline, between- and within-person MAP, between- and within-person BMI, between- and within-person alcohol consumption, baseline age, sex, baseline smoking pack-years, PASE score, LDL cholesterol, HDL cholesterol, and HbA1c. Interactions with follow-up time were included to accommodate the estimation of time dependent effects for between-person MAP and BMI, average alcohol consumption, baseline age, sex, smoking pack-years, PASE score, LDL cholesterol, HDL cholesterol, and HbA1c.^19^ The modelling of time interactions allowed assessment of whether these characteristics were associated with differences in the longitudinal trajectory of each retinal vascular feature. Such modelling of time interactions in longitudinal studies is key to study the impact of variables on dependent variable trajectories.^51^ Within-person terms were entered without interactions with follow-up time, and represent associations between within-person variation in the exposure and concurrent retinal vascular variation.

Models were developed using MixedLM (Statsmodels, Python)^52^. The models were fitted using restricted maximum likelihood (REML) and the L-BFGS method, with a maximum of 3,000 iterations.

### Retinal vascular change and subsequent clinical outcomes

Retinal vascular features have been associated with risk of cardiovascular disease and all-cause death.^5,8,39,53^ In this study, we sought to determine whether the extent of change between retinal vascular traits over time is associated with increased risk of cardiovascular disease or death.

Cox proportional hazards models were fitted using follow-up 1 as a landmark to investigate subsequent all-cause mortality and major adverse cardiovascular events (MACE). Whilst numerous definitions of MACE exist, we elected to utilise a three-point definition, which was highlighted as amongst the most common MACE definitions in a previous literature review.^22^ Accordingly, MACE was defined as myocardial infarction, stroke, or cardiovascular death (defined as death caused by stroke or heart disease). Participants with the outcome before or at follow-up 1 were excluded from the corresponding analysis, and only events occurring after follow-up 1 were considered incident events. Participants who refused to answer or answered ‘don’t know’ to age of disease were excluded. The CLSA recorded the date at which the participants’ status was last known, and this date was used for censoring, as well as using non-cardiovascular death as a censoring event in the case of MACE.

Cox models were fitted separately for each retinal vascular metric. Changes in the retinal vascular metric and its baseline value were independently standardised within each complete-case analysis sample, such that hazard ratios (HR) represent the association with a one-SD increase in baseline-to-follow-up 1 change. Models additionally included sex, with female sex as the reference category, the baseline value of the corresponding retinal vascular metric, and the interval in years between the baseline and follow-up 1 data collection site visits.

Attained age was used as the underlying time scale^21^. Participants entered the risk set at their age at follow-up 1 and exited at their age at the event or censoring. The use of attained age as a time scale in Cox models has been asserted as the most flexible control for age effects and avoids the need to include age as a covariate.^21^ Participants were necessarily required to have complete data for the retinal change metric (i.e. a good quality baseline and follow-up 1 colour fundus photograph) in order to be included in the analysis. Event status was ascertained at follow-up 2 or via the decedent questionnaire.

### Multiple testing correction

Benjamini-Hochberg false discovery rate (FDR) correction was used for multiple testing correction (significance threshold of p < 0.05 after correction).^54^ For Cox proportional hazards, FDR was applied separately for each clinical outcome across the retinal vascular metrics tested. For Pearson correlation of vascular metric change scores, FDR was applied across all unique trait combinations (minus correlations along the diagonal where *x*=*y*). For linear mixed models, correction was applied for each trait across the vascular metrics tested.

### Software and versions

All analyses were completed in Python 3.10.6. Data were organised using Pandas 1.5.1 and Numpy 1.23.4. Plots were constructed using Seaborn 0.13.2.^55^ Correlation analyses and multiple testing corrections were conducted using SciPy 1.13.1.^49^ Linear mixed models were fitted using Statsmodels 0.13.5.^52^ Cox proportional-hazards models were fitted using Lifelines 0.27.4.^56^

## Code availability

All code, used to analyse this data, minus the AutoMorph pipeline which has its own repository (https://github.com/rmaphoh/AutoMorph.git), is presented in this GitHub repository: https://github.com/TomJulian/CLSA-Vascular-Longitudinal-Associations.git

## Data availability statement

Data are available from the Canadian Longitudinal Study on Aging (www.clsa-elcv.ca) for researchers who meet the criteria for access to de-identified CLSA data.

## Acknowledgements

We acknowledge the following sources of funding: the Medical Research Council (MRC) Clinical Research Training Fellowship (MR/Z504105/1, to T.H.J.) and MR/W006804/1 to G.R.M; the Wellcome Trust (224643/Z/21/Z, Clinical Research Career Development Fellowship, to P.I.S.); the Royal Academy of Engineering under the RAEng Chair in Emerging Technologies (INSILEX CiET1919\/19), ERC Advanced Grant-UKRI Frontier Research Guarantee (INSILICO EP\/Y030494/1), the UK Centre of Excellence on In-Silico Regulatory Science and Innovation (UK CEiRSI) (10139527) and the BHF Manchester Centre of Research Excellence (RE\/24/130017), to A.F.F.; and the National Institute for Health and Care Research (NIHR) Manchester Biomedical Research Centre (BRC) (NIHR203308) to A.F.F., J.H., H.D., J.D., P.I.S. and T.H.J. B.K. acknowledges support from the British Heart Foundation Research Excellence Award (RE/24/130017) and BHF Personal Chair (CH/13/2/30154).

This research was made possible using the data/biospecimens collected by the Canadian Longitudinal Study on Aging (CLSA). Funding for the Canadian Longitudinal Study on Aging (CLSA) is provided by the Government of Canada through the Canadian Institutes of Health Research (CIHR) under grant reference: LSA 94473 and the Canada Foundation for Innovation, as well as the following provinces, Newfoundland, Nova Scotia, Quebec, Ontario, Manitoba, Alberta, and British Columbia. This research has been conducted using Retinal Scans (Baseline), Retinal Scans (FUP1), Retinal scans (FUP2), Comprehensive Baseline v7.2, Comprehensive Follow-up 1 v5.1, Comprehensive Follow-up 2 v4.2, Decedent questionnaire version 2.0, Participant status version 5.0.under Application ID 2407009. The CLSA is led by Drs. Parminder Raina, Christina Wolfson and Susan Kirkland. The time and commitment of the participants to the CLSA study platform is gratefully acknowledged, without whom this research would not be possible.The opinions expressed in this manuscript are the author’s own and do not necessarily reflect the views of the Canadian Longitudinal Study on Aging.

---

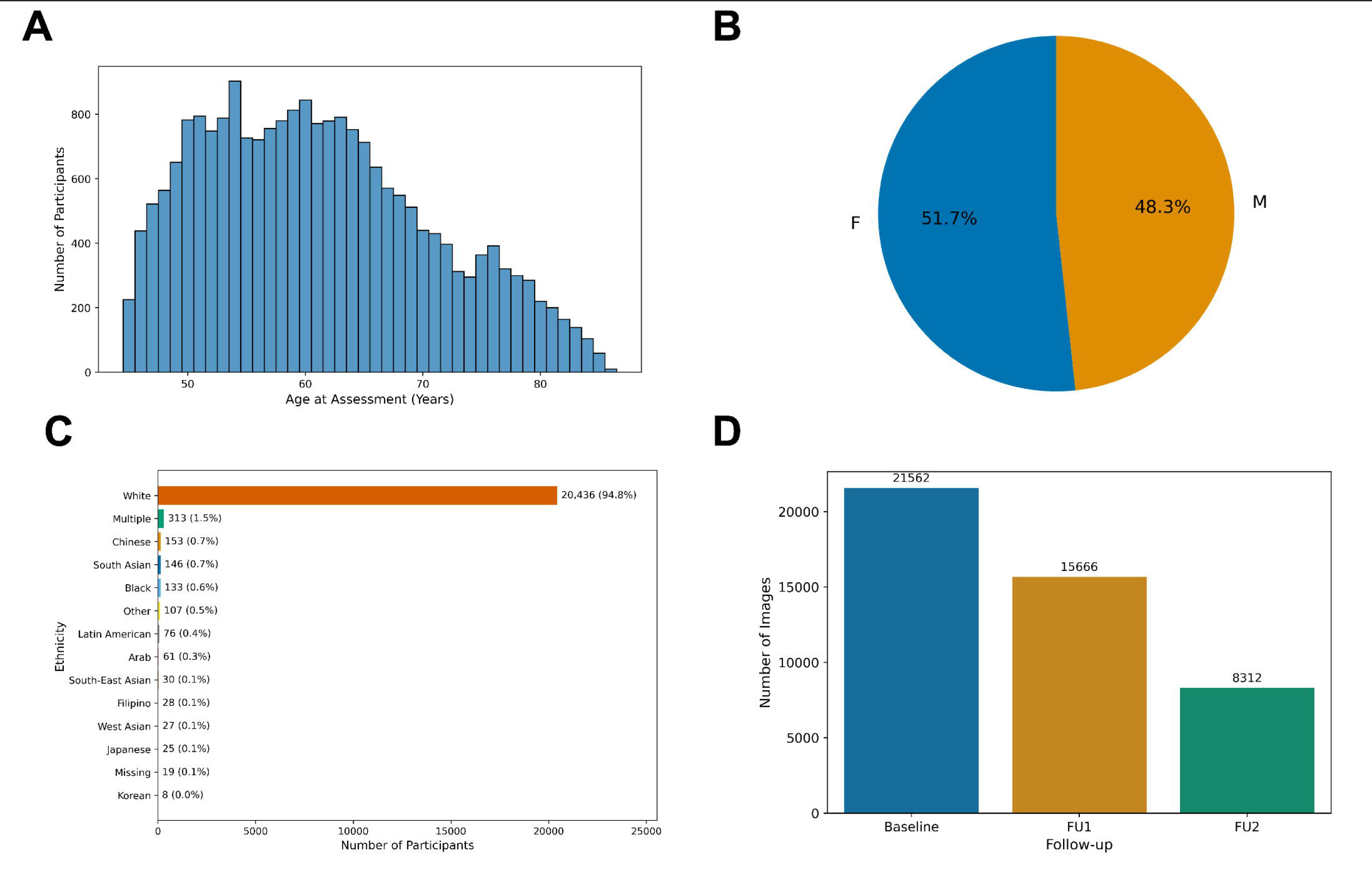

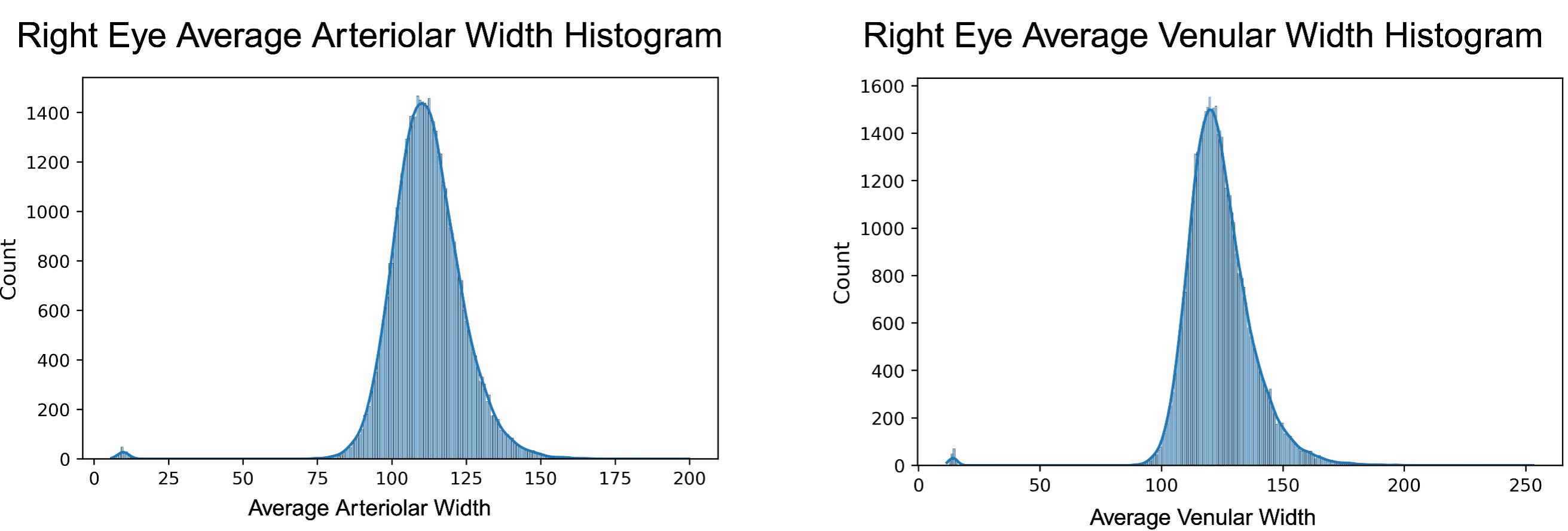

